# Forecasting COVID-19 pandemic in Poland according to government regulations and people behavior

**DOI:** 10.1101/2020.05.26.20112458

**Authors:** Magdalena Orzechowska, Andrzej K. Bednarek

## Abstract

The coronavirus disease 2019 (COVID-19) outbreak is a worldwide pandemic problem that started in China in December 2019 and within a few months spread to all continents. Very high infectivity of SARS-CoV-2 virus and substantial disease severity caused medical care capacity shortage in many countries. Therefore, real-time epidemic forecasting of the COVID-19 is useful to plan public health strategies like country lockdown and healthcare reorganization.

We used extended susceptible-infected-removed (eSIR) model to predict the epidemic trend of COVID-19 in Poland under different scenarios of the lockdown and lockdown removal. We used time-series data of SARS-CoV-2 infection from March 4 to May 22 2020. Our forecast includes the impact of a timeline of preventive measures introduced in Poland. Using eSIR algorithm we estimated the basic reproductive number and a total number of infections under different epidemic trend scenarios.

Using eSIR modeling we estimated that the basic reproductive number in Poland concerning different scenarios of the lockdown removal is in a range of 3.91-4.79. The lowest predicted number of infected cases would be 263 900 (0 - 1 734 200, 95%CI) if the strict protective measures were maintained until the end of September. However, under different scenarios of precautions removal, a total number of infected cases may exceed one million within the next year.

Relatively early introduction of strong precautions in Poland significantly slowed down epidemic spread in Poland in comparison with other European countries like Italy or Spain. However, early removal of protective measures may result in a significant increase in infection. Data shows that the number of new COVID-19 cases in Poland beyond May 18 is linear what could be a prognosis of a duration of the epidemic exceeding 300 days.

## 1 Introduction

The global pandemic of Corona Virus Disease 2019 (COVID-19) spread over the majority of the human population all over the world. It started as a small local outbreak in Wuhan, China end of 2019 but shortly became the major health problem in the whole of China. Within the first months of 2020 COVID-19 has spread to whole Asia, Europe and the Americas with an important number of cases reported in Africa and Australia. Shortly in some countries, a huge scale of cases has forced governments to introduce large restrictions on people-to-people contacts by closing schools, public transport, shops, services and factories, and also by closing state borders. On May 22, 2020, above 5×10^6 cases were reported globally, with 1.6×10^6 in the US, above 300×10^3 in Brazil and Russia, and above 200×10^3 in several European countries (UK, Spain, Italy). Interestingly, on May 22 the total case number in China was around 85×10^3 (source JHU-CRC). The very high infectivity of coronavirus SARS-CoV-2 is accompanied by a severe course of the disease in a significant number of patients. As a result, in many countries, there have been problems in accessing hospital medical care and deficiencies in life-saving equipment such as respirators. Global deaths number associated with COVID-19 exceeded 340×10^3 on May 22 (source JHU-CRC). In many countries, governments introduced various regulations aimed at slowing down and limiting the spread of disease. The most restrictive regulations appeared, among others, in China and Italy. It was the closure of cities and regions, where the incidence was the highest, and the order to stay at home. Due to such severe restrictions covering virtually all residents, proper and accurate monitoring of the spread of the virus is very important. It is very difficult to maintain such a regime for a long time so prognosis of epidemics time points is crucial to maintain readiness for medical assistance and to plan an effective control strategy. Epidemiologists use several algorithms to forecast the course of the epidemic, like SIR compartment model [1], which serve as a base for several modified model used for COVID-19 forecasting. The discrete-time SIR model including dead individuals was used for prognosis the course of the epidemic in Hubei, China [2]. The modified susceptible-exposed-infected-removed (SEIR) epidemiological model was used to forecast the outbreak in Wuhan, China [3]. Casella used a new mathematical model based on SIR which includes control strategies and test reporting delay [4]. Another new outbreak modeling strategy was proposed by Giordano et al. SIDARTHE (**S**usceptible, **I**nfected, **D**iagnosed, **A**iling, **R**ecognized, **T**hreatened, **H**ealed, **E**xtinct). This algorithm, used for Italy outbreak forecasting, discriminates between diagnosed and non-diagnosed infected individuals which is very important for assessing the spread of the virus since quarantined persons do not participate in the further spread of the disease [5]. All those models used no or very little input of social and business isolation strategies, which forced on a large scale have a very significant impact on the spread of the epidemics. Wangping et al. compared COVID-19 outbreaks in Italy with Hunan, China according to the time-line of introducing subsequent government regulations and self-increasing self-awareness [6]. They used extended SIR model (eSIR) previously used in forecasting outbreak in China [7]. This algorithm is easily adaptable to different factors modifying COVID-19 spread. Despite mentioned regulations ordered by the government, quarantine and immunization factors can be included in the calculation. Based on differences in government-ordered regulations and recommendations to limit the rate of SARS-CoV-2 spread, Wangping et al. concluded that such big difference between Italy and Hunan is based on initially less restrictive regulations and a few days delay in their implementation [6].

In this report, we show forecast models of COVID-19 outbreak in Poland concerning various strategies for abolishing restrictive regulations. The first laboratory-confirmed SARS-CoV-2 infected person was reported in Poland on March 4, 2020 [8]. On March 12 the Polish government introduced the first package of regulations aimed at stopping the epidemic rate (closing restaurants, pubs and bars, movie theaters, closing schools, university classes and canceling mass events). However, on March 12 there were already 51 laboratory-confirmed cases, the first COVID-19 death and only 2234 tests performed in total [8]. In the following weeks’ further orders and recommendations were introduced: March 16 Poland closed its land borders and international airports for passenger traffic; March 25 limiting non-family gatherings to two people and religious gatherings to six and forbidding non-essential travel, closing parks, boulevards, beaches, hairdressers and beauty salons, forbidding unaccompanied minors from exiting their homes; April 16 making it obligatory to cover one’s nose and mouth in public places [8]. On April 16 Poland had 7918 laboratory-confirmed cases with average 10×10^A^6 tests performed daily. Starting May 18 Polish government declared to loosen restrictions allowing more passengers in public transport and markets, re-opening pubs and restaurants with guests number limitation and opening hairdressers and beauty salons with some precautionary measures as well as allowing outdoor activities [8]. Unfortunately, many citizens took it as offsetting the threat of infection, they stopped wearing masks and reduced social distance. By using eSIR modeling we build several forecast models depending on loosening restrictions and recommendations as well as people behavior.

## 2 Materials and Methods

### 2.1 Retrieval of the epidemiological data

In the present study, we retrieved the publicly available epidemiological data on COVID-19 prevalence in Poland of the time frame including the very beginning of the Polish spread of COVID-19, 4 March 2020 till 22 May 2020, provided by the government in real-time that include the cumulative number of confirmed and recovered cases as well as deaths from COVID-19 [9].

The employed data are related to unidentified patients, collected and officially published to the public by the Polish authorities, thereby no ethical approval was required to conduct the study.

### 2.2 The forecast of COVID-19 spread in Poland with a time-varying transmission rate – the model applied

The trend of COVID-19 transmission was estimated through the R0, a reproduction number reflecting the transmissibility of a virus that spreads in an uncontrolled manner, which is defined as the average number of new infections generated by each infected person [10]. The R0≤1indicates the trend towards decline and eventual disappearance of COVID-19.

To forecast the spread of the virus in Poland according to the government regulations and civilians behavior, we calculated the R0 by applying an extended state-specific SIR (eSIR) epidemiological model with a time-varying transmission rate π(t) that incorporates forms of social and medical isolation (e.g. government isolation protocols, personal protective measures, community-level isolation, environmental changes etc.) within the infectious disease dynamic system that is available as eSIR package dedicated for R environment [7]. In greater detail, the eSIR model bases upon daily-updated time series of infected and removed (either recovered or dead) cumulative as an input as well as Markov Chain Monte Carlo (MCMC) algorithm, thus enables the assessment of the effectiveness of social isolation protocols for confining COVID-19 spread across the specific region (herein: Poland). Both predicted turning points and their credible bands of the epidemiological trend of COVID-19 may be obtained from the eSIR under a given quarantine protocol. The model includes three major turning points, which are defined as follows: the first is the mean predicted time when the daily number of infected cases drops down below the previous ones; the second is the mean predicted time when the daily number of removed cases exceeds the number of infected cases, and the third, i.e. the end-point, is the time when the median of currently infected cases turns to zero. All figures demonstrating the computed forecast with i.a. spaghetti plot are generated automatically by the eSIR package.

In our study, we used the input data for Poland that come from 2020 March 4 to 2020 May 22 this year. We specified four potential models of various transmission rate modifiers π(t) with accordance to actual interventions in different times of COVID-19 spread by analogy to the Chinese and Italian studies [6]. Figure 2 shows time-lines of Polish preventive measures starting March 4 until May 18 and since May 18 four predicted transmission rate π(t) changes according to the withdrawal of restrictions models and drop in self-awareness. Regarding the government isolation protocols and measures, we set π(t) = 0.95 if t ∊ (Mar 4, Mar 12), no government-ordered isolation, some level of the self-awareness; π(t) = 0.7 if t ∊ (Mar 12, Mar 16), closing schools, university classes and canceling mass events; π(t) = 0.5 if t ∊ (Mar 16, Mar 25), Poland closed its land borders and international airports for passenger traffic, allowing only Polish citizens/residents and those with immediate Polish family to enter the country. All people entering Poland were subjected to the compulsory 14 days quarantine; π(t) = 0.4 if t ∊ (Mar 25, Apr 16), limiting non-family gatherings to two people and religious gatherings to six as well as forbidding non-essential travels across the country, closing parks, boulevards, beaches, beauty salons, and unaccompanied minors from leaving their homes; π(t) = 0.2 if t ∊ (Apr 16, May 18), obligatory covering one’s mouth and nose in the public areas and respective π(t) = 0.2, π(t) = 0.4, π(t) = 0.6, π(t) = 0.8 if t > May 18, representing four different models of the visible drop in caution and gradual departing from the restrictions. As a comparison, a model assuming no precautionary measures has been computed (π(t) =1). Moreover, as recommended by the authors, we set M=5e5 and nburnin=2e5 to obtain stable MCMC chains. All analyses were performed using R version 4.0.0. The code enabling conducting eSIR forecast for Poland with the input data is available as Supplementary File 1.

## 3 Results

Referring to the official statistics of COVID-19 in Poland on 22 May 2020, the cumulative number of new cases, deaths and recovers was estimated as a total of 20615, 993 and 8731, respectively (Figure 1). As observed, the cumulative number of infected is relatively low but a daily number of new cases is a flat line with a slight slope increase after May 18.

**Figure 1.**
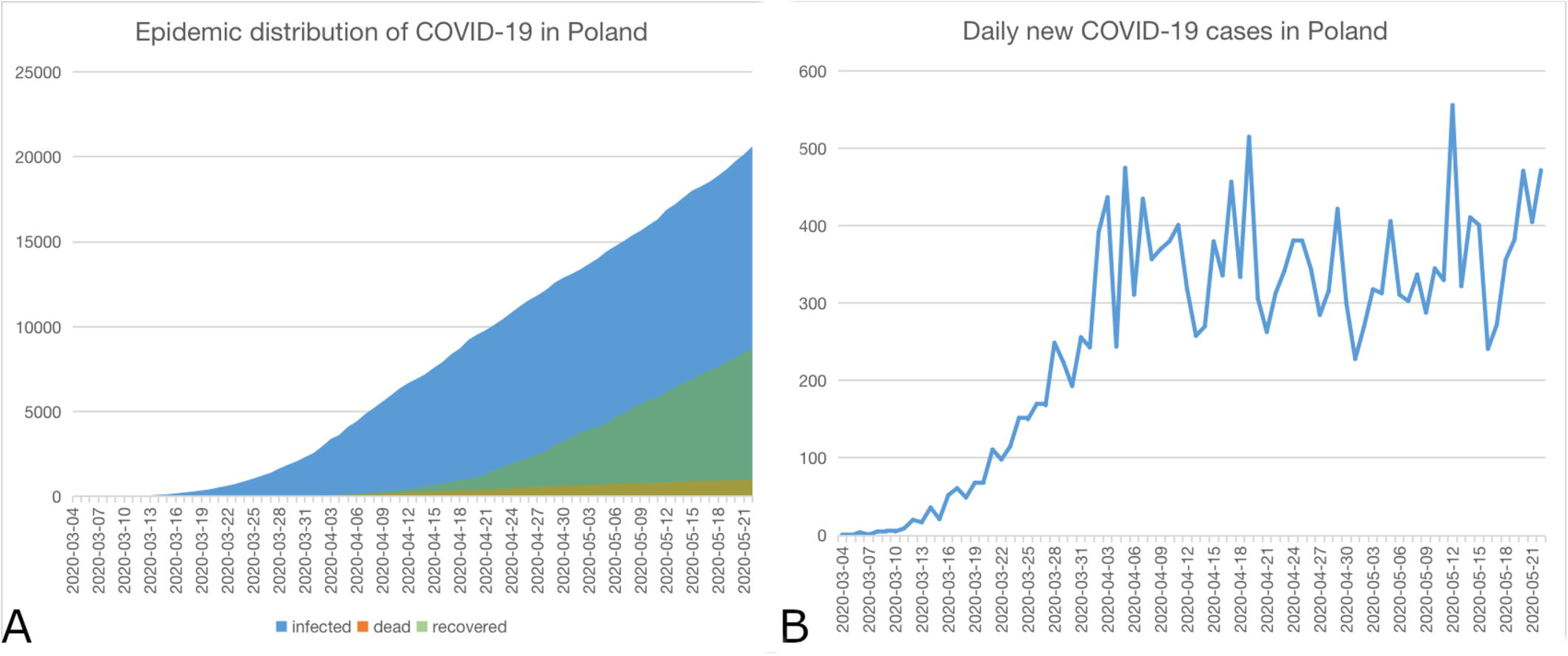
COVID-19 spread in Poland. A: epidemic distribution; B: new cases daily.

The first preventive measures against COVID-19 were introduced in Poland on March 12, following by additional regulations on March 16, March 25, April 16 as described above (Figure 2).

**Figure 2.**
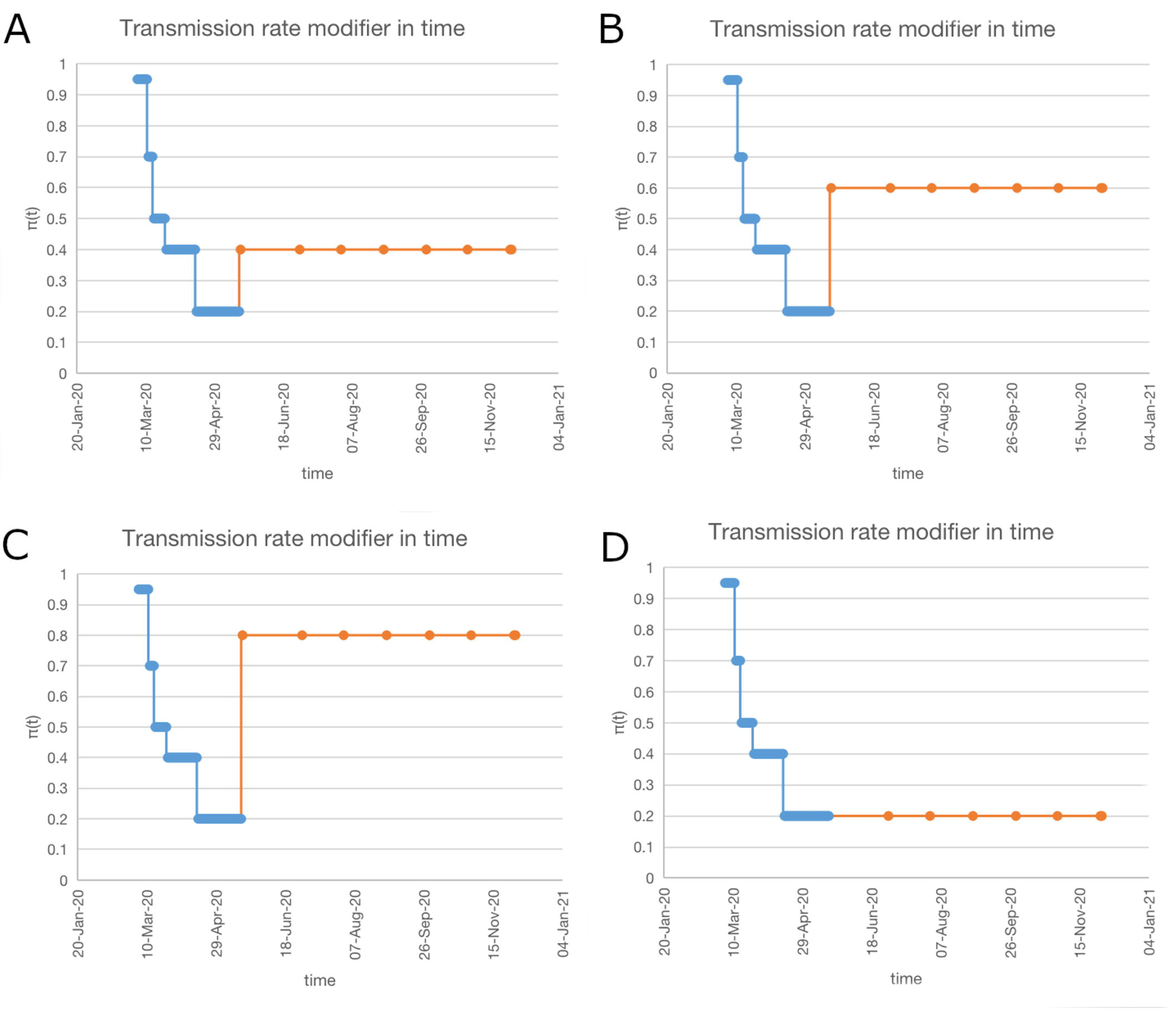
Timeline of introduction of preventive measures and their effect on time-varying transmission rates: π(t) set to 0.95; 0.7; 0.5; 0.4 and 0.2 on day March 4; March 12; March 16; March 25 and April 16 respectively. A-C assumption of π(t) (0.4; 0.6; 0.8) effective since May 18 depending on the lifting restrictions and people behavior. D: hypothetical π(t) = 0.2 prolonged for an indefinite period beyond May 18.

According to the eSIR model of COVID-19 prevalence in Poland, the estimated posterior mean values of R0 depends on a type and time-line of preventive measures withdrawn. Table 1 shows the estimated R0 values and epidemic endpoints according to different epidemic models in Poland.

**Table 1:** Detailed statistics of eSIR forecast for Poland involving different variants of loosening the preventive restrictions.

| $\pi(t)$ | R0 | | | $\beta$ | | endpoint | |
| --- | --- | --- | --- | --- | --- | --- | --- |
|  | median | mean | 95%CI | mean | 95%CI | mean | 95%CI |
| 1.00 | 2.81 | 2.93 | 1.59-4.97 | 0.37 | 0.06-1.00 | 2020/06/16 | 2020/03/14-inf |
| 0.20 | 4.70 | 4.79 | 2.54-7.56 | 0.17 | 0.07-0.30 | 2020/11/12 | 2020/11/07-inf |
| 0.40 | 4.53 | 4.61 | 2.49-7.18 | 0.16 | 0.07-0.28 | 2020/12/23 | 2020/09/22-inf |
| 0.60 | 4.20 | 4.28 | 2.35-6.68 | 0.15 | 0.06-0.25 | inf | inf-inf |
| 0.80 | 3.91 | 3.99 | 2.24-6.19 | 0.13 | 0.06-0.22 | inf | inf-inf |

Figure 3 shows forecasts of epidemiological trends of COVID-19 in Poland under different introduced preventions and its predicted withdrawal. As may be seen in Figure 3E and Table 1 in a case of now any preventive measures epidemic would spread in Poland very fast. The β factor (the expected number of people an infected person infects per day) is significantly high without any prevention. In such a case epidemic endpoint will come on June 16 and mean of 54% population infected (up to 99%; 95%CI) on the end of June. On another hand, precautions measures until December 2020 (π(t) = 0.2) will end up with only 0.7% (260×10^3 people) infected within a very prolonged time. According to different predicted effects of prevention lifting, all the assumed time-varying transmission rates show a similar effect with above 1 million people infected in a year or more.

**Figure 3.**
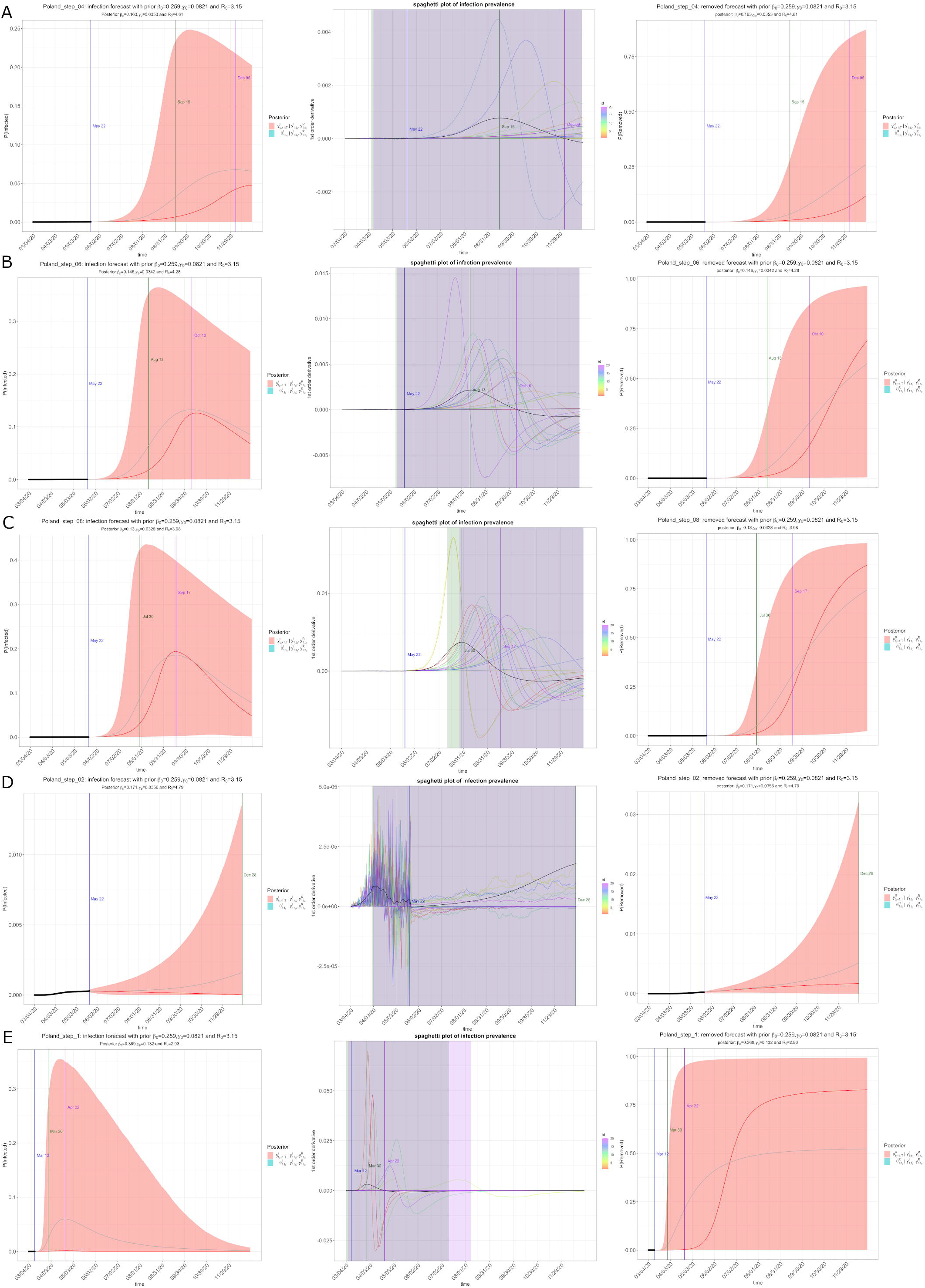
Forecasts of COVID-19 epidemic under different effects of preventive measures. A-C: predicted epidemiological trend according to changes in time-varying transmission rates; A: π(t) = 0.4; B: π(t) = 0.6; C: π(t) = 0.8. D and E are hypothetical epidemiological trends in case of D: maintaining strict prevention, π(t) = 0.2 and E: no any precautions introduced, π(t) = 1.0. A-E: on the left - forecast of a daily number of cases as a percentage of the population; on the right - forecast of a daily number of recoveries as a percentage of the population; in the middle - spaghetti plot, in which randomly selected MCMC draws of the first-order derivative of the posterior prevalence of infection.

## 4 Discussion

Starting end of 2019 a novel strain of coronavirus SARS-CoV-2 originated in Wuhan, China and spread globally. The first confirmed cases were identified in December 2019 [11]. On January 30 infection was found in 19 countries and 7 818 cases were reported [12]. On March 11 WHO declared the pandemic [13]. Then, it was 118 319 confirmed cases and 4292 deaths [14]. Although most cases show no symptoms, the very high rate of infections in several countries resulted in a shortage of healthcare capacity due to a small number of beds equipped with respirators and oxygen as well as high morbidity among medical personnel [15]. Therefore many governments decided to introduce lockdown to slow-down COVID-19 spread. China decided to block the cities and to introduce strict quarantine for all residents of highly affected regions. The other preventive measures were: isolation of infected and suspected of being infected, social distancing. Such methods have proven effective in slowing down the spread of the virus and gave additional time for medical care adaptation [16]. The first two most affected countries were China and Italy. Several reports are forecasting how coronavirus infection rate is affected by preventive measures [17,18]. However, there is a limited number of publications considering timing in preventive regulations [5-7,19]. The most advanced model seems to be eSIR [6]. In the study comparing outbreak in Hunan, China and Italy eSIR model explains differences in the rate of infection, clearly identifying that 1-7 days delay in the country lockdown would result in even 5 times more infections [5]. As a consequence, the capacity for healthcare may be insufficient and the disease may spread to medical staff and store employees, which will further increase the extent of the epidemic. Therefore the introduction of preventive measures, even very broad and restrictive may prevent a population from health disaster and an elevated number of death cases as a consequence of a shortage in medical equipment. The question is how long such intensive preventive measures should last and when we can depart from strict social isolation, allow business activities and initiate a return to normal functioning. This is a negotiating factor between SARS-CoV-2 spread prevention and economic protection of people and the state. There is no country which may sustain months of lockdown without a threat of deep recession. Based on eSIR model, to prevent a substantial growth of COVID-19 cases Italy should continue lockdown by August 5.

We performed eSIR analysis for Poland using different scenarios of loosening lockdown regulations. The first preventive measures were introduced in Poland on March 12, a week after the first laboratory-confirmed case of COVID-19, when the outbreak was already spread in other European countries, with the particularly dangerous way in Italy. The timetable of a stepped lockdown in Poland is shown in Figure 2. Our model shows that preventive measures slowed down coronavirus spread in Poland in Figure 3 and Table 1. As shown, introducing and maintaining restrictive lockdown slows down the spread of the epidemic but the final effect depends on how long specific preventive measures will continue. On May 18 some precautions have been lifted and according to social observations, Polish change their behavior to further reduce the social distance. Our forecast shows that keeping preventive measures like before May 18 would keep epidemic under strict control with relatively low infectivity per day with the epidemic endpoint in December. However, starting May 18 we estimate that π(t) values will rise to 0.4, 0.6 or 0.8 depending on the social distance reduction resulting from changes in regulations and people’s behaviour. In the event of a significant reduction of precautions, our model shows a very significant increase in a daily number of infections with the peak falling between the end of September and December (Figure 3A-C) and more than 1 million people infected within one year. A significant local increase in the number of new cases was observed in the Silesia region due to coronavirus spread among miners [9]. This shows that some economic sectors and employee groups are particularly vulnerable because of the difficulty or even the inability to observe strict precautions. Our model shows that even slight lifting of preventive measures in Poland starting middle of May will result in an increase of the frequency of infection with peak time on the end of September or later (Figure 3).

Our forecast model was built on limited data availability; on May 22 in Poland was 719 571 tests performed in total since February 29, which is roughly 2% of the population tested. Spatial distribution of infected cases shows the possibility of a local outbreak model. However, there is no spatially fine-grained surveillance data for model tuning. Our model does not include the incubation period and the duration of infectivity. R0 in our forecast for Poland is high and exceeds 4 but is similar to that in Italy according to eSIR model, where R0 is adjusted according to the timetable of prevention [5]. On another hand, our calculated R0 in Poland may be overestimated due to the relatively low number of tests, which were mainly carried out on suspected people.

In conclusion, to our knowledge, this is the first model forecasting COVID-19 spread in Poland concerning the time-line of introduced preventive measures and its anticipated stepped withdrawn. The forecast shows that relatively early introduction of preventive measures in Poland caused a significant reduction in the epidemic rate in comparison with Italy or Spain. However, the future depends mostly on people behavior which is the most difficult factor to control. Loss of self-awareness and a significant reduction in social distance may result in a sharp increase in the daily incidence of SARS-CoV-2 infections and a shortage in a medical care capacity. Our model suggests that relatively rigorous precautions should be maintained in Poland until the end of September to keep the epidemic rate low.

## Data Availability

The authors confirm that the data supporting the findings of this study are available within the article and its supplementary materials. These data were derived from the public domain.

https://www.gov.pl/web/koronawirus

https://www.gov.pl/web/koronawirus/wykaz-zarazen-koronawirusem-sars-cov-2

## Data Availability Statement

The datasets analyzed for this study can be found in the public domain: https://www.gov.pl/web/koronawirus and https://www.gov.pl/web/koronawirus/wykaz-zarazen-koronawirusem-sars-cov-2.

## Supplementary Data

**Supplementary File 1**. R code for eSIR forecast for Poland including the input data.

